# A national cohort study of COVID-19 in-hospital mortality in South Africa: the intersection of communicable and non-communicable chronic diseases in a high HIV prevalence setting

**DOI:** 10.1101/2020.12.21.20248409

**Authors:** Waasila Jassat, Cheryl Cohen, Stefano Tempia, Maureen Masha, Susan Goldstein, Tendesayi Kufa, Pelagia Murangandi, Dana Savulescu, Sibongile Walaza, Jamy-Lee Bam, Mary-Ann Davies, Hans W. Prozesky, Jonathan Naude, Ayanda T. Mnguni, Charlene A. Lawrence, Hlengani T. Mathema, Jarrod Zamparini, John Black, Ruchika Mehta, Arifa Parker, Perpetual Chikobvu, Halima Dawood, Ntshengedzeni Muvhango, Riaan Strydom, Tsholofelo Adelekan, Bhekizizwe Mdlovu, Nirvasha Moodley, Eunice L. Namavhandu, Paul Rheeder, Jacqueline Venturas, Nombulelo Magula, DATCOV author group, Lucille Blumberg

**Affiliations:** National Institute for Communicable Disease of the National Health Laboratory Service, Johannesburg, South Africa; Influenza Division, Centers for Disease Control and Prevention, Atlanta, Georgia, United States of America; School of Public Health, Faculty of Health Sciences, University of the Witwatersrand, Johannesburg, South Africa; MassGenics, Duluth, Georgia, United States of America; Right to Care, Johannesburg, South Africa; South Africa Medical Research Council (SAMRC) Centre for Health Economics and Decision Science-PRICELESS SA, School of Public Health, University of the Witwatersrand, Johannesburg, South Africa; Division of Global HIV & TB, Centers for Disease Control and Prevention, Pretoria, South Africa; Western Cape Department of Health, Cape Town, South Africa; Tygerberg Hospital and Division of Infectious Disease, University of Stellenbosch, Cape Town, South Africa; Mitchells Plain District Hospital, Cape Town, South Africa; Khayelitsha District Hospital, Cape Town, South Africa; Department of Medicine, Charlotte Maxeke Johannesburg Academic Hospital and the University of the Witwatersrand, Johannesburg, South Africa; Walter Sisulu University, Livingstone Hospital, Nelson Mandela Bay, South Africa; Klerksdorp-Tshepong Hospital and the University of Witwatersrand, Klerksdorp, South Africa; Free State Department of Health, Bloemfontein, South Africa; Grey’s Hospital, Pietermaritzburg, South Africa; Limpopo Department of Health, Polokwane, South Africa; Northern Cape Department of Health, Kimberley, South Africa; Gauteng Department of Health, Johannesburg, South Africa; Mpumalanga Department of Health, Nelspruit, South Africa; KwaZulu-Natal Department of Health, Pietermaritzburg, South Africa; Eastern Cape Department of Health, Bisho, South Africa; University of Pretoria, Pretoria, South Africa; University of KwaZulu-Natal, Durban, South Africa

**Keywords:** COVID-19, hospital admissions, HIV, comorbid disease, mortality

## Abstract

**Background:** The interaction between COVID-19, non-communicable diseases, and chronic infectious diseases such as HIV and tuberculosis (TB) are unclear, particularly in low- and middle-income countries in Africa. South Africa has a national adult HIV prevalence of 19% and TB prevalence of 0.7%. Using a nationally representative hospital surveillance system in South Africa, we investigated the factors associated with in-hospital mortality among individuals with COVID-19.

**Methods:** Using data from national active hospital surveillance, we describe the demographic characteristics, clinical features, and in-hospital mortality among hospitalised individuals testing positive for SARS-CoV-2, during 5 March 2020 to 27 March 2021. Chained equation multiple imputation was used to account for missing data and random effect multivariable logistic regression models were used to assess the role of HIV-status and underlying comorbidities on in-hospital COVID-19 mortality.

**Findings:** Among the 219,265 individuals admitted with laboratory confirmed SARS-Cov-2, 51,037 (23.3%) died. Most commonly observed comorbidities among individuals with available data were hypertension (61,098/163,350; 37.4%), diabetes (43,885/159,932; 27.4%), and HIV (13,793/151,779; %), while TB was reported in 3.6% (5,282/146,381) of individuals. While age was the most important predictor, other factors associated with in-hospital COVID-19 mortality were HIV infection [aOR 1.34, 95% CI: 1.27-1.43), past TB [aOR 1.26, 95% CI: 1.15-1.38), current TB [aOR 1.42, 95% CI: 1.22-1.64) and both past and current TB [aOR 1.48, 95% CI: 1.32-1.67) compared to never TB, as well as other described risk factors for COVID-19, such as male sex, non-white race, and chronic underlying hypertension, diabetes, chronic cardiac disease, chronic renal disease, and malignancy. After adjusting for other factors, PLWH not on ART [aOR 1.45, 95% CI: 1.22-1.72] were more likely to die in-hospital compared to PLWH on ART. Among PLWH, the prevalence of other comorbidities was 29.2% compared to 30.8% among HIV-uninfected individuals. Increasing number of comorbidities was associated with increased mortality risk in both PLWH and HIV-uninfected individuals.

**Interpretation:** Identified high risk individuals (older individuals and those with chronic comorbidities and PLWH, particularly those not on ART) would benefit from COVID-19 prevention programmes such as vaccine prioritisation, as well as early referral and treatment.

**Funding:** South African National Government

**Research in context:** *Evidence before this study:* Since the emergence of the COVID-19 pandemic, studies have identified older age, male sex and presence of underlying comorbidities including heart disease and diabetes as risk factors for severe disease and death. There are very few studies, however, carried out in low- and middle-income countries (LMIC) in Africa, many of whom have high poverty rates, limited access to healthcare, and high prevalence of chronic communicable diseases, such as HIV and tuberculosis (TB). Data are also limited from settings with limited access to HIV treatment programmes. Early small cohort studies mainly from high income countries were not conclusive on whether HIV or TB are risk factors for disease severity and death in COVID-19 patients. Large population cohort studies from South Africa’s Western Cape province and the United Kingdom (UK) have found people living with HIV (PLWH) to have a moderately increased risk of COVID-19 associated mortality. Of these, only the Western Cape study presented data on mortality risk associated with presence of high viral load or immunosuppression, and found similar levels of severity irrespective of these factors. Recent meta-analyses have confirmed the association of HIV with COVID-19 mortality. No studies reported on the interaction between HIV-infection and other non-communicable comorbidities on COVID-19 associated mortality. We performed separate literature searches on PubMed using the following terms: “COVID-19” “risk factors” and “mortality”; “HIV” “COVID-19” and “mortality”; “TB” “COVID-19” and “mortality”. All searches included publications from December 1, 2019 until May 5, 2021, without language restrictions. Pooled together, we identified 2,786 published papers. Additionally, we performed two literature searches on MedRxiv using the terms “HIV” “COVID-19” and “mortality”, and “TB” “COVID-19” and “mortality” from April 25, 2020 until May 5, 2021, without language restrictions. Pooled together, we identified 7,744 pre-prints.

*Added value of this study:* Among a large national cohort of almost 220,000 individuals hospitalised with COVID-19 in a setting with 19% adult HIV prevalence and 0.7%TB prevalence, we found that along with age, sex and other comorbidities, HIV and TB were associated with a moderately increased risk of in-hospital mortality. We found increasing risk of in-hospital mortality among PLWH not on ART compared to those on ART. Among PLWH, the prevalence of other comorbidities was high (29%) and the effect of increasing numbers of comorbidities on mortality was similar in PLWH and HIV-uninfected individuals. Our study included 13,793 PLWH from all provinces in the country with varying levels of access to HIV treatment programmes.

*Implications of all the available evidence:* The evidence suggests that PLWH and TB-infected individuals should be prioritised for COVID-19 prevention and treatment programmes, particularly those with additional comorbidities. Increasing age and presence of chronic underlying illness are important additional factors associated with COVID-19 mortality in a middle-income African setting. The completeness of data is a limitation of this national surveillance system, and additional data are needed to confirm these findings.

## Introduction

The first case of COVID-19 was documented in South Africa on 5 March 2020. By 27 March 2021, 1,545,431 cases had been reported and the cumulative incidence was 2,592.0 cases per 100,000 persons. (1) Meta-analyses have reported in-hospital mortality rates between 15% and 24%, with substantial global and temporal heterogeneity. (2-4) Worldwide, studies have linked COVID-19 mortality to older age, male sex, and underlying medical conditions, including cardiac disease, diabetes, cancer, chronic pulmonary disease, obesity, and kidney disease. (2,3,5,6) Race or ethnicity and poverty were associated with increased risk of death in COVID-19 cases in large population cohorts and meta-analyses. (7-9)

Characterizing populations at increased risk of COVID-19 mortality is important for prioritization of interventions, particularly in low- and middle-income countries (LMIC) where resources are limited. However, available data on risk factors for severe COVID-19 disease, including mortality, are mostly from high-income countries (HIC). High poverty rates, limited access to healthcare and high prevalence of chronic communicable diseases such as HIV and TB, likely affects the burden of COVID-19 and disease severity in LMICs. (10)

South Africa is a LMIC country with coinciding epidemics of non-communicable diseases (NCD) and chronic infectious diseases (HIV and TB). In 2020, the prevalence of HIV in the adult population 15-49 years was 19%, (11) and TB prevalence 0.7%. (12) In 2019, there were 7.5 million people estimated to be living with HIV in South Africa, of which 2.3 million (31%) were eligible but not receiving treatment. (13)

Understanding whether HIV and TB are associated with mortality among individuals hospitalised with COVID-19 is of critical importance for South Africa and other countries in the region with large HIV and/or TB epidemics, in order to guide public health action around prevention and treatment of COVID-19. Early single centre cohort (14,15) studies and meta-analyses (16,17) from HICs with relatively small numbers of people living with HIV (PLWH) did not find HIV to be a risk factor for severe COVID-19 disease. Larger population cohorts (18-20) and more recent meta-analyses (21-23) found PLWH to have an increased population risk of COVID-associated mortality. The interaction between other comorbidities, HIV infection, and COVID-19 in-hospital mortality is not well described.

In this paper, we examine the association between HIV infection, other comorbidities and in-hospital mortality among hospitalised individuals with laboratory-confirmed SARS-CoV-2 using data from a national surveillance programme in South Africa from 5 March 2020 to 27 March 2021.

## Methods

### Study setting and data sources

South Africa is administratively divided into nine provinces. South Africa has a dual health system with a publicly funded district health system, that serves approximately 84% of the population, and a private health system largely funded by private health insurance schemes. (24)

In the absence of existing national COVID-19 hospital surveillance systems, the National Institute for Communicable Diseases (NICD) established DATCOV as a national surveillance system for COVID-19 hospitalizations on 1 April 2020. (25) DATCOV was adopted for national implementation following South African government endorsement on 15 July 2020, and by October 2021 full coverage of all hospitals had been achieved. By 27 March 2021, a total of 393 public-sector hospitals and 251 private-sector hospitals have reported COVID-19 hospitalisations on DATCOV. Further details about the DATCOV surveillance system including its implementation, data management and data quality assurance are described in supplementary methods (page 1, Supplementary Material).

### Potential risk factors and covariates

Age, sex, race, and comorbidities (hypertension, diabetes, chronic cardiac disease, asthma, other chronic respiratory disease, chronic renal disease, malignancy in the past five years, HIV and past and current TB) were considered as potential risk factors for COVID-19 in-hospital mortality. Data on comorbidities including HIV, ART status, and CD4 and viral load (VL) done in the past 12 months, were submitted to DATCOV by the hospital, based on information contained in the patient’s written hospital record, and were not independently verified by the DATCOV team and neither were laboratory records accessed to obtain this data. Date of most recent CD4 or VL was not documented. The level of virological control or immunosuppression was assessed based on available VL result within the past year and categorised as virologically suppressed (HIV-RNA<1,000 copies/ml) or as viraemic (HIV-RNA≥1,000 copies/ml); and immune reconstituted (CD4 count≥200 cells/μl) or immunosuppressed (CD4 count<200 cells/μl). (26) CD4 count is no longer routinely performed on all patients, as per South African HIV care guidelines. (27)

In-hospital mortality was defined as a death related to COVID-19 that occurred during the hospital stay and excluded deaths that occurred due to other causes or after discharge from hospital. Because the main outcome of the study was in-hospital mortality, all analyses were implemented among SARS-Cov-2 hospitalised patients with a known in-hospital outcome (i.e. discharged alive or died) at the time of data extraction (27 March 2021).

### Statistical analysis

We analysed DATCOV data from 5 March 2020 to 27 March 2021. For the main analysis, to account for incomplete or missing data on selected variables, we used multivariate imputation by chained equation (MICE) and generated ten complete imputed datasets that were used for subsequent analyses. Variables analysed using MICE included sex, race, month of admission and comorbidities including HIV and TB infection, hypertension, diabetes, asthma, malignancy, and chronic pulmonary, cardiac and renal diseases. ART, HIV viral load and CD4 counts were also incomplete and were conditionally imputed only among HIV-positive patients (either with observed or imputed HIV status). Complete variables included in the imputation process were age, province, health sector (i.e. public or private) and in-hospital outcome (i.e. discharged alive or died). Descriptive statistics such as frequencies and percentages were used for categorical variables, and continuous variables were expressed as mean with standard deviation (SD) or median and interquartile range (IQR) on the imputed datasets.

We implemented post-imputation random effect (on admission facility) multivariable logistic regression models to: (i) compare PLWH and individuals who were HIV-uninfected; (ii) determine the factors associated with in-hospital mortality, including ART, HIV viral load and CD4 count among HIV-positive individuals; and (iii) evaluate the combined effect of multiple non-HIV comorbidities on COVID-19 in hospital mortality. A combined comorbidity variable categorised absence, one, two, and three or more comorbidities among those that were individually significantly associated with in-hospital mortality in model (ii) above. We evaluated the effect of multiple comorbidities among any patient and among PLWH and HIV-uninfected patients separately through stratification by HIV infection status. In addition, we assessed potential differential effect (through the inclusion of an interaction term) of multiple or individual comorbidities on in-hospital mortality between PLWH and HIV-uninfected individuals.

A random effect on admission facility was included for all analyses to account for potential differences in the service population and the quality of care at each facility. For each multivariable model we assessed all variables that were significant at p<0.2 on univariate analysis (to evaluate lack of significance at the univariate analysis after adjusting for potential confounders) and dropped non-significant factors (p≥0.05) with manual backward elimination. Pairwise interactions were assessed by inclusion of product terms for all variables remaining in the final multivariable additive model. We implemented several sensitivity analyses separately analysing risk factors for mortality in the public and private sector using post-imputation random effect multivariable logistic regression models. We also reported the univariate and multivariate association of all covariates evaluated in the analyses described above to the main outcomes (i.e. HIV infection or in-hospital mortality) using non-imputed data. The statistical analysis was implemented using Stata 15 (Stata Corp®, College Station, Texas, USA). We followed STROBE guideline recommendations.

### Ethical considerations

The Human Research Ethics Committee (Medical), University of the Witwatersrand, approved the project protocol as part of a national surveillance program (M160667). In South Africa, surveillance for notifiable medical conditions such as COVID-19 requires health facilities to submit data on all cases of emerging pathogens to national authorities. As such, individual consent for inclusion of their data to DATCOV is waivered. All personal identifying information was de-linked for our analysis and stored in a secure server.

### Role of the funding source

DATCOV as a national surveillance system, is funded by the NICD and the South African National Government. No additional funding was obtained towards the completion of this analysis and the development of this manuscript. The funders of the study had no role in study design, data collection, data analysis, data interpretation, or writing of the report. The corresponding author had full access to all the data in the study and had final responsibility for the decision to submit for publication.

## Results

### Study population

From 5 March 2020 to 27 March 2021, 229,154 individuals were hospitalised with laboratory-confirmed SARS-Cov-2 and reported to DATCOV. Of these, 219,265 met the inclusion criteria (Figure S1, page 4 Supplementary Materials). The percent of missing data for the imputed variables varied and was highest for race (33.5%), and for HIV-specific variables including HIV diagnosis (30.8%), being on ART (41.4%), viral load (87.6%) and CD4 count (79.9%) (Table S1, page 5 Supplementary Materials).

South Africa experienced a first wave of SARS-Cov-2 infection, peaking at 10,086 admissions per week in July 2020, and a second wave peaking at 17,976 admissions per week in January 2021 (Figure S2, page 7 Supplementary Materials).

The median age of SARS-Cov-2 hospitalised patients was 54 years (interquartile range [IQR] 40 – 66) and 55.7% of patients (121,937/218,827) were female (Table S1, page 5 Supplementary Materials). Race information was available for 145,876 (66.5%) patients, of whom 114,571 (78.5%) were black. The public sector accounted for 51.9% (113,856/219,265) of hospitalisations reported to DATCOV. Most admissions (176,272/219,265 [80.4%]) were recorded by hospitals in four provinces, namely Western Cape, Gauteng, KwaZulu-Natal and Eastern Cape.

Following multiple imputation, the estimated proportion of individuals reporting at least one comorbidity was 60.5% (95%CI: 59.8%-61.2%). The most prevalent comorbidities were hypertension (38.6%), diabetes (29.0%), and HIV (11.7%). The prevalence of non-communicable comorbidities increased with age, while HIV and TB were most prevalent in individuals aged 20–59 years (Figure 3).

### Factors associated with HIV infection

Of 151,779 (69.2%) hospitalised patients with available data on HIV, 13,793 (9.1%) were PLWH (Table 1). The HIV prevalence was 20.4% in the public sector and was 2.2% in the private sector (age-specific HIV prevalence for public and private sector presented in Table S2, page 8 Supplementary Materials). A subset of the 13,793 PLWH had ART (8078 [58.6%]), viral load (1,716 [12.4%]) and CD4 (2,770 [20.1%]) information available. Of these, 92.7% (7,484/8,078) were receiving ART, 25.8% (443/1,716) were viraemic (HIV-RNA≥1000 copies/ml) and 39.0% (1,080/2,770) were immunosuppressed (CD4 count<200 cells/μl) based on last available test.

**Table 1:**
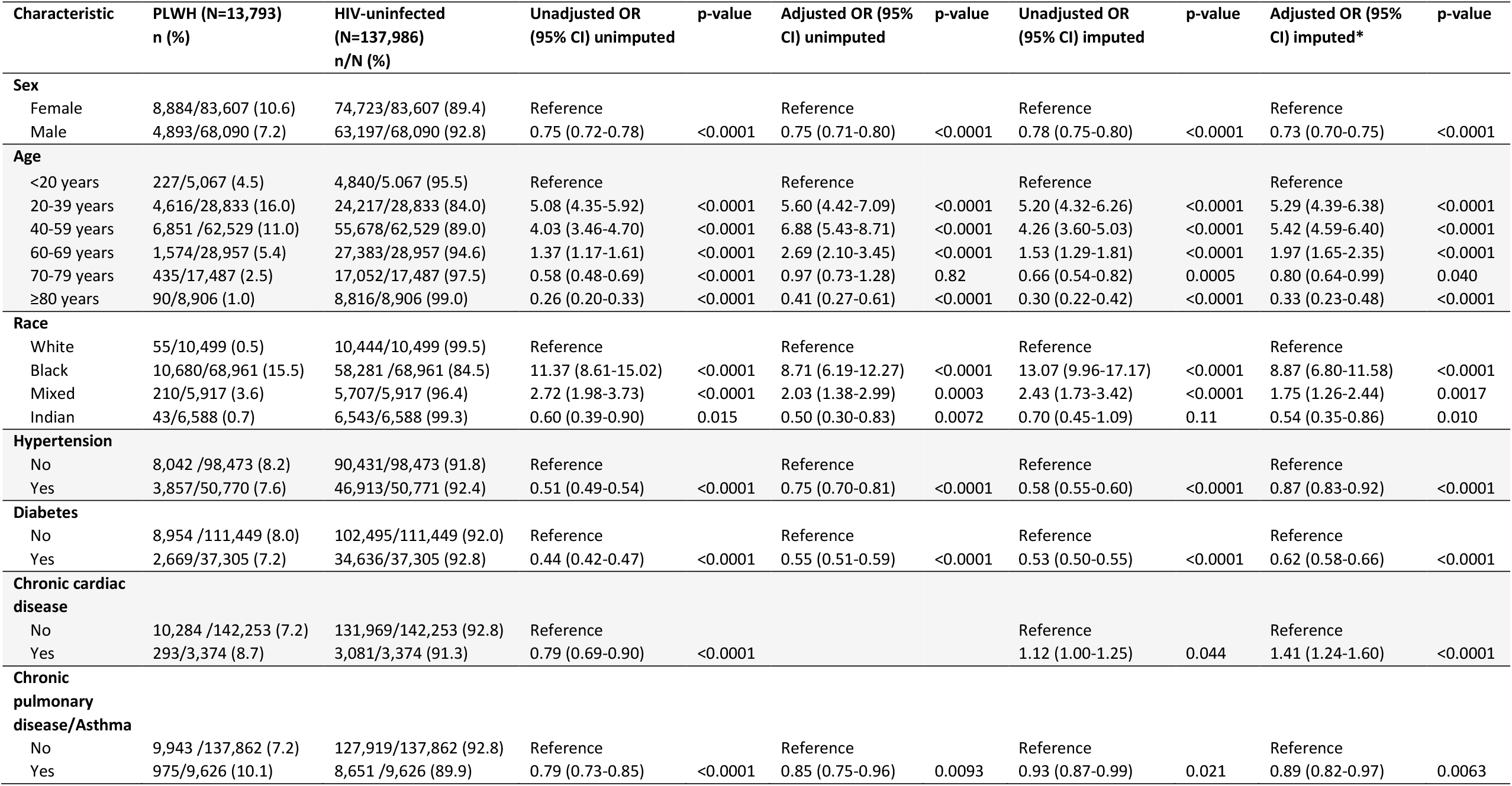
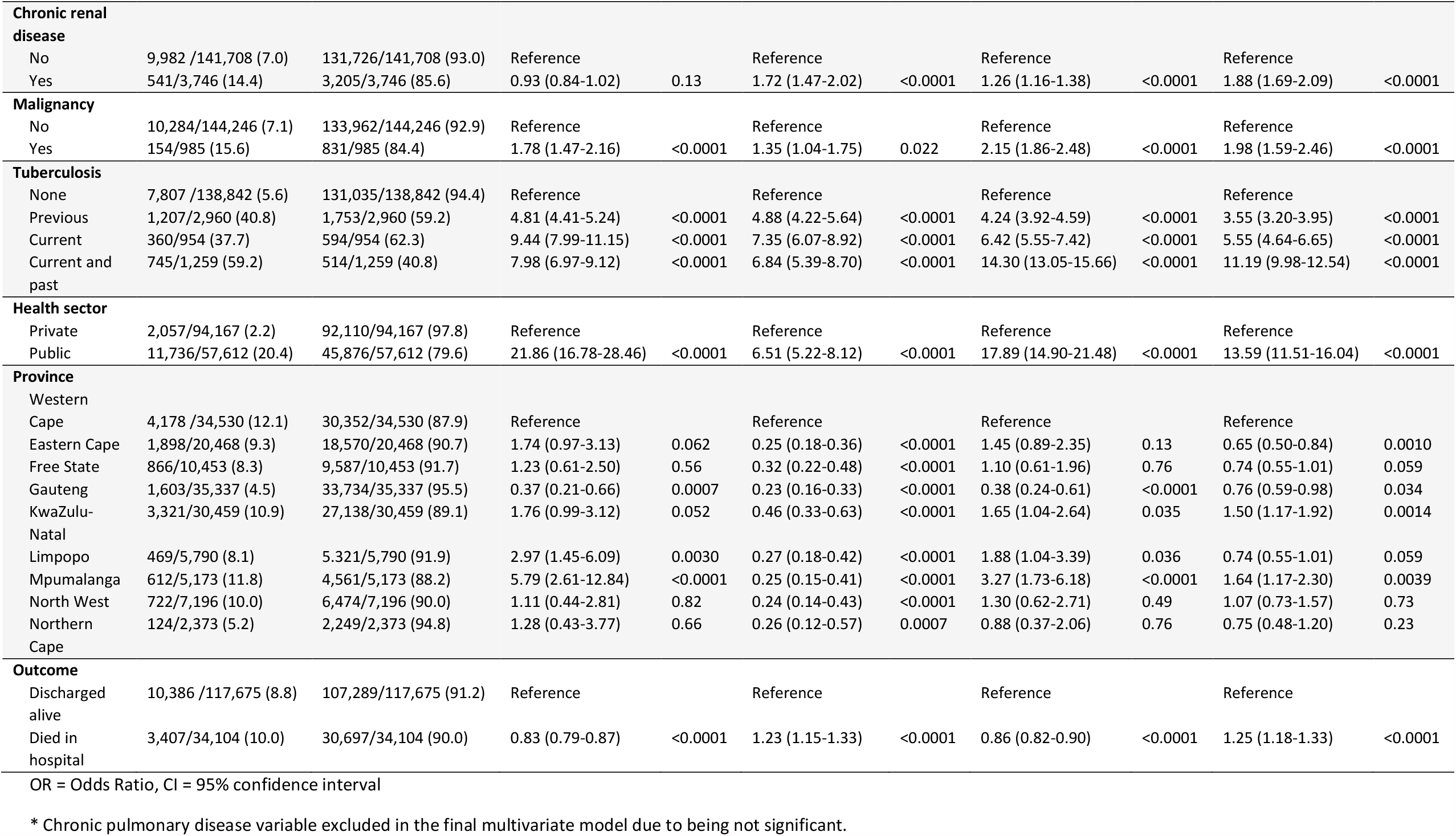
Characteristics of PLWH and HIV-uninfected individuals with laboratory-confirmed SARS-CoV-2 admitted to hospital, 5 March 2020-27 March 2021, DATCOV, South Africa (N=219,265)

Compared to HIV-uninfected SARS-CoV-2 hospitalised patients, PLWH were more likely to be aged 20-39 years, 40-59 years, and 60-69 years, compared to <20 years; black or mixed race compared to white race; admitted in the public health sector; to have comorbid chronic cardiac disease, renal disease, malignancy, current TB, past TB and previous and current TB, and to die in hospital. PLWH were less likely to be aged 70-79 years and ≥80 years; male; of Indian ancestry; and to have comorbid hypertension, diabetes, and chronic pulmonary disease or asthma. HIV prevalence of COVID-19 patients varied with lower prevalence in Eastern Cape and Gauteng province, and higher prevalence in KwaZulu-Natal and Mpumalanga provinces (Table 1 and Figure 2).

### In-hospital COVID-19-associated mortality

Of the 219,265 hospitalised patients with recorded outcomes, 168,228 (76.7%) were discharged alive and 51,037 (23.3%) died. The unadjusted in-hospital case-fatality ratio (CFR) for PLWH was (3,407/13,793 [24.7%]) compared to (30,697/137,986 [22.3%]) for HIV-uninfected individuals.

Factors statistically associated with in-hospital COVID-19 mortality were increasing age 20-39 years, 40-59 years, 60-69 years, 70-79 years and ≥80 years, compared to <20 years; male sex; black, mixed race, and Indian ancestry compared to white race; having comorbid hypertension, diabetes, chronic cardiac disease, chronic renal disease, malignancy, HIV, past TB, current TB or both past and current TB, and being admitted in the public health sector. In-hospital mortality increased each month of the epidemic to the peak of the first wave in July then decreased between waves and increased again to the peak of the second wave in January. In-hospital mortality was significantly higher in five provinces, Eastern Cape, Free State, KwaZulu-Natal, Limpopo and Mpumalanga, compared to the Western Cape (Table 2 and Figure 3). The un-imputed multivariate analysis showed similar findings (Table 2).

**Table 2:**
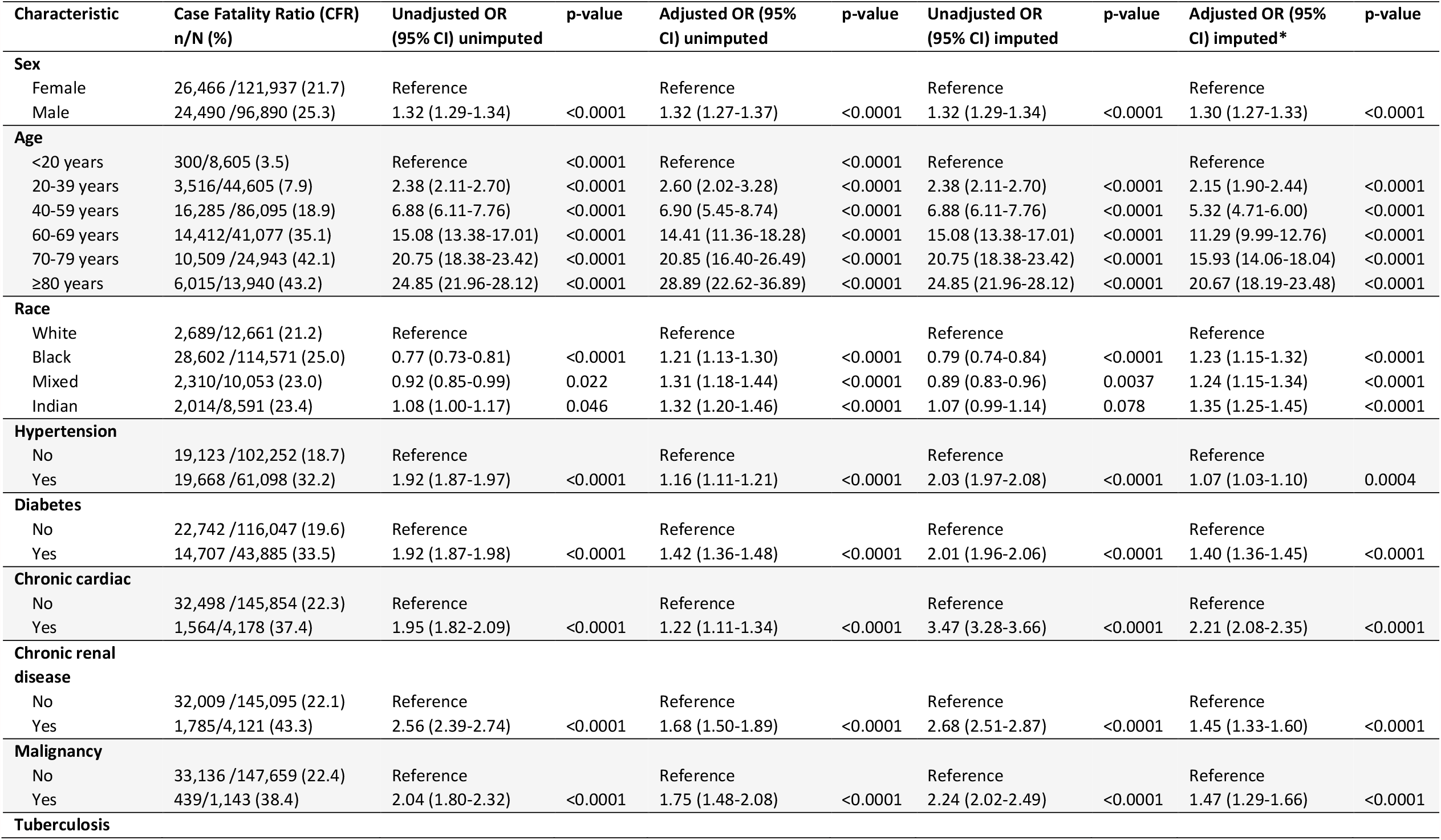
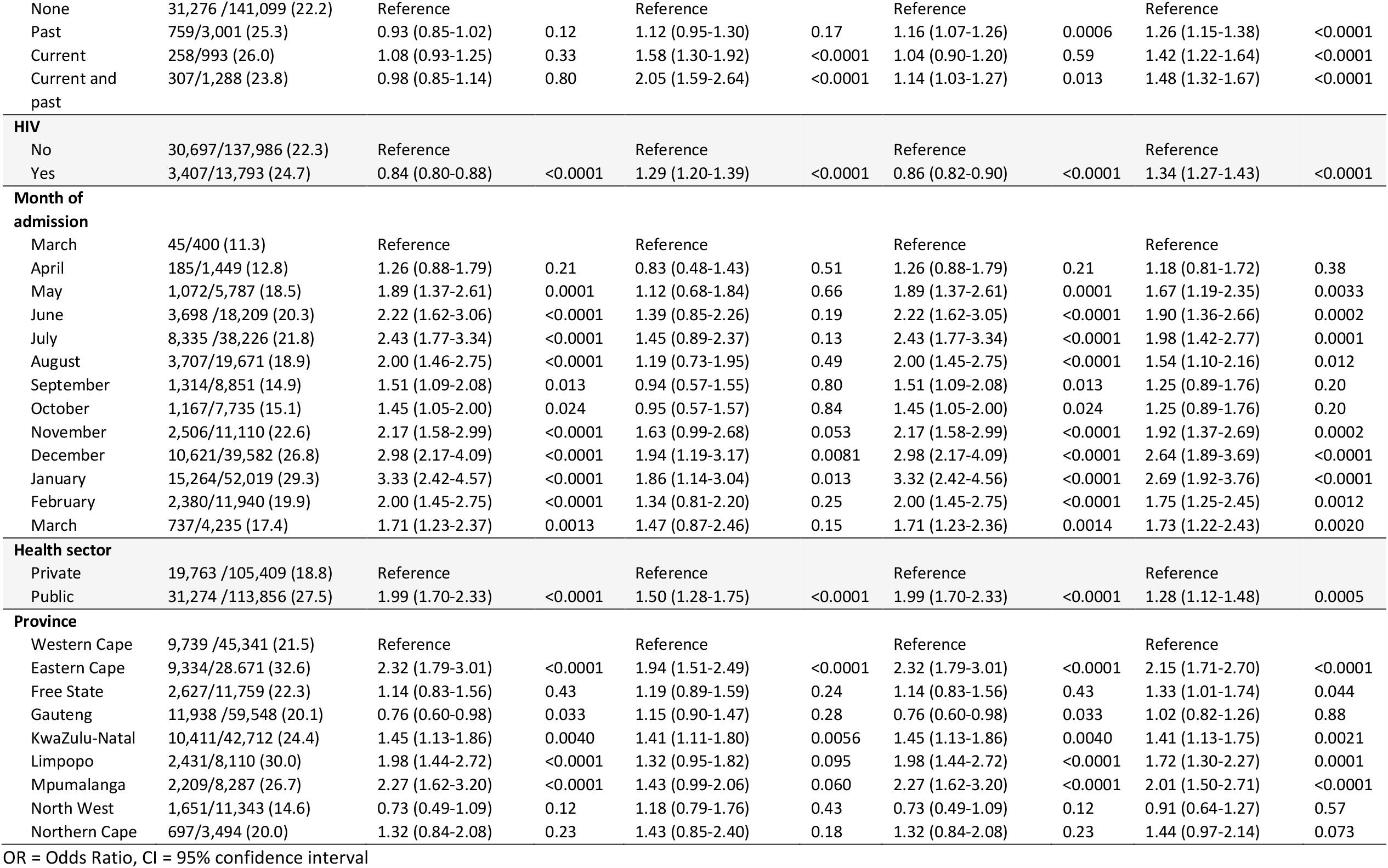
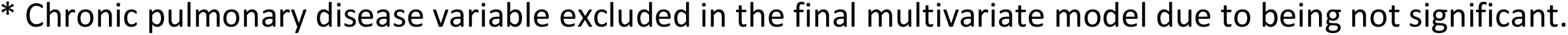
Factors associated with in-hospital mortality among individuals with laboratory-confirmed SARS-CoV-2 admitted to hospital, 5 March 2020-27 March 2021, DATCOV, South Africa. (N=219,265)

| Characteristic | Case Fatality Ratio (CFR)<br>n/N (%) | Unadjusted OR<br>(95% CI) unimputed | p-value | Adjusted OR (95%<br>CI) unimputed | p-value | Unadjusted OR<br>(95% CI) imputed | p-value | Adjusted OR (95%<br>CI) imputed* | p-value |
| --- | --- | --- | --- | --- | --- | --- | --- | --- | --- |
| **Sex** |  |  |  |  |  |  |  |  |  |
| Female | 26,466 /121,937 (21.7) | Reference |  | Reference |  | Reference |  | Reference |  |
| Male | 24,490 /96,890 (25.3) | 1.32 (1.29-1.34) | <0.0001 | 1.32 (1.27-1.37) | <0.0001 | 1.32 (1.29-1.34) | <0.0001 | 1.30 (1.27-1.33) | <0.0001 |
| **Age** |  |  |  |  |  |  |  |  |  |
| <20 years | 300/8,605 (3.5) | Reference | <0.0001 | Reference | <0.0001 | Reference |  | Reference |  |
| 20-39 years | 3,516/44,605 (7.9) | 2.38 (2.11-2.70) | <0.0001 | 2.60 (2.02-3.28) | <0.0001 | 2.38 (2.11-2.70) | <0.0001 | 2.15 (1.90-2.44) | <0.0001 |
| 40-59 years | 16,285 /86,095 (18.9) | 6.88 (6.11-7.76) | <0.0001 | 6.90 (5.45-8.74) | <0.0001 | 6.88 (6.11-7.76) | <0.0001 | 5.32 (4.71-6.00) | <0.0001 |
| 60-69 years | 14,412/41,077 (35.1) | 15.08 (13.38-17.01) | <0.0001 | 14.41 (11.36-18.28) | <0.0001 | 15.08 (13.38-17.01) | <0.0001 | 11.29 (9.99-12.76) | <0.0001 |
| 70-79 years | 10,509 /24,943 (42.1) | 20.75 (18.38-23.42) | <0.0001 | 20.85 (16.40-26.49) | <0.0001 | 20.75 (18.38-23.42) | <0.0001 | 15.93 (14.06-18.04) | <0.0001 |
| ≥80 years | 6,015/13,940 (43.2) | 24.85 (21.96-28.12) | <0.0001 | 28.89 (22.62-36.89) | <0.0001 | 24.85 (21.96-28.12) | <0.0001 | 20.67 (18.19-23.48) | <0.0001 |
| **Race** |  |  |  |  |  |  |  |  |  |
| White | 2,689/12,661 (21.2) | Reference |  | Reference |  | Reference |  | Reference |  |
| Black | 28,602 /114,571 (25.0) | 0.77 (0.73-0.81) | <0.0001 | 1.21 (1.13-1.30) | <0.0001 | 0.79 (0.74-0.84) | <0.0001 | 1.23 (1.15-1.32) | <0.0001 |
| Mixed | 2,310/10,053 (23.0) | 0.92 (0.85-0.99) | 0.022 | 1.31 (1.18-1.44) | <0.0001 | 0.89 (0.83-0.96) | 0.0037 | 1.24 (1.15-1.34) | <0.0001 |
| Indian | 2,014/8,591 (23.4) | 1.08 (1.00-1.17) | 0.046 | 1.32 (1.20-1.46) | <0.0001 | 1.07 (0.99-1.14) | 0.078 | 1.35 (1.25-1.45) | <0.0001 |
| **Hypertension** |  |  |  |  |  |  |  |  |  |
| No | 19,123 /102,252 (18.7) | Reference |  | Reference |  | Reference |  | Reference |  |
| Yes | 19,668 /61,098 (32.2) | 1.92 (1.87-1.97) | <0.0001 | 1.16 (1.11-1.21) | <0.0001 | 2.03 (1.97-2.08) | <0.0001 | 1.07 (1.03-1.10) | 0.0004 |
| **Diabetes** |  |  |  |  |  |  |  |  |  |
| No | 22,742 /116,047 (19.6) | Reference |  | Reference |  | Reference |  | Reference |  |
| Yes | 14,707 /43,885 (33.5) | 1.92 (1.87-1.98) | <0.0001 | 1.42 (1.36-1.48) | <0.0001 | 2.01 (1.96-2.06) | <0.0001 | 1.40 (1.36-1.45) | <0.0001 |
| **Chronic cardiac** |  |  |  |  |  |  |  |  |  |
| No | 32,498 /145,854 (22.3) | Reference |  | Reference |  | Reference |  | Reference |  |
| Yes | 1,564/4,178 (37.4) | 1.95 (1.82-2.09) | <0.0001 | 1.22 (1.11-1.34) | <0.0001 | 3.47 (3.28-3.66) | <0.0001 | 2.21 (2.08-2.35) | <0.0001 |
| **Chronic renal disease** |  |  |  |  |  |  |  |  |  |
| No | 32,009 /145,095 (22.1) | Reference |  | Reference |  | Reference |  | Reference |  |
| Yes | 1,785/4,121 (43.3) | 2.56 (2.39-2.74) | <0.0001 | 1.68 (1.50-1.89) | <0.0001 | 2.68 (2.51-2.87) | <0.0001 | 1.45 (1.33-1.60) | <0.0001 |
| **Malignancy** |  |  |  |  |  |  |  |  |  |
| No | 33,136 /147,659 (22.4) | Reference |  | Reference |  | Reference |  | Reference |  |
| Yes | 439/1,143 (38.4) | 2.04 (1.80-2.32) | <0.0001 | 1.75 (1.48-2.08) | <0.0001 | 2.24 (2.02-2.49) | <0.0001 | 1.47 (1.29-1.66) | <0.0001 |
| **Tuberculosis** |  |  |  |  |  |  |  |  |  |
| None | 31,276 /141,099 (22.2) | Reference |  | Reference |  | Reference |  | Reference |  |
| Past | 759/3,001 (25.3) | 0.93 (0.85-1.02) | 0.12 | 1.12 (0.95-1.30) | 0.17 | 1.16 (1.07-1.26) | 0.0006 | 1.26 (1.15-1.38) | <0.0001 |
| Current | 258/993 (26.0) | 1.08 (0.93-1.25) | 0.33 | 1.58 (1.30-1.92) | <0.0001 | 1.04 (0.90-1.20) | 0.59 | 1.42 (1.22-1.64) | <0.0001 |
| Current and past | 307/1,288 (23.8) | 0.98 (0.85-1.14) | 0.80 | 2.05 (1.59-2.64) | <0.0001 | 1.14 (1.03-1.27) | 0.013 | 1.48 (1.32-1.67) | <0.0001 |
| **HIV** |  |  |  |  |  |  |  |  |  |
| No | 30,697/137,986 (22.3) | Reference |  | Reference |  | Reference |  | Reference |  |
| Yes | 3,407/13,793 (24.7) | 0.84 (0.80-0.88) | <0.0001 | 1.29 (1.20-1.39) | <0.0001 | 0.86 (0.82-0.90) | <0.0001 | 1.34 (1.27-1.43) | <0.0001 |
| **Month of admission** |  |  |  |  |  |  |  |  |  |
| March | 45/400 (11.3) | Reference |  | Reference |  | Reference |  | Reference |  |
| April | 185/1,449 (12.8) | 1.26 (0.88-1.79) | 0.21 | 0.83 (0.48-1.43) | 0.51 | 1.26 (0.88-1.79) | 0.21 | 1.18 (0.81-1.72) | 0.38 |
| May | 1,072/5,787 (18.5) | 1.89 (1.37-2.61) | 0.0001 | 1.12 (0.68-1.84) | 0.66 | 1.89 (1.37-2.61) | 0.0001 | 1.67 (1.19-2.35) | 0.0033 |
| June | 3,698 /18,209 (20.3) | 2.22 (1.62-3.06) | <0.0001 | 1.39 (0.85-2.26) | 0.19 | 2.22 (1.62-3.05) | <0.0001 | 1.90 (1.36-2.66) | 0.0002 |
| July | 8,335 /38,226 (21.8) | 2.43 (1.77-3.34) | <0.0001 | 1.45 (0.89-2.37) | 0.13 | 2.43 (1.77-3.34) | <0.0001 | 1.98 (1.42-2.77) | 0.0001 |
| August | 3,707/19,671 (18.9) | 2.00 (1.46-2.75) | <0.0001 | 1.19 (0.73-1.95) | 0.49 | 2.00 (1.45-2.75) | <0.0001 | 1.54 (1.10-2.16) | 0.012 |
| September | 1,314/8,851 (14.9) | 1.51 (1.09-2.08) | 0.013 | 0.94 (0.57-1.55) | 0.80 | 1.51 (1.09-2.08) | 0.013 | 1.25 (0.89-1.76) | 0.20 |
| October | 1,167/7,735 (15.1) | 1.45 (1.05-2.00) | 0.024 | 0.95 (0.57-1.57) | 0.84 | 1.45 (1.05-2.00) | 0.024 | 1.25 (0.89-1.76) | 0.20 |
| November | 2,506/11,110 (22.6) | 2.17 (1.58-2.99) | <0.0001 | 1.63 (0.99-2.68) | 0.053 | 2.17 (1.58-2.99) | <0.0001 | 1.92 (1.37-2.69) | 0.0002 |
| December | 10,621/39,582 (26.8) | 2.98 (2.17-4.09) | <0.0001 | 1.94 (1.19-3.17) | 0.0081 | 2.98 (2.17-4.09) | <0.0001 | 2.64 (1.89-3.69) | <0.0001 |
| January | 15,264/52,019 (29.3) | 3.33 (2.42-4.57) | <0.0001 | 1.86 (1.14-3.04) | 0.013 | 3.32 (2.42-4.56) | <0.0001 | 2.69 (1.92-3.76) | <0.0001 |
| February | 2,380/11,940 (19.9) | 2.00 (1.45-2.75) | <0.0001 | 1.34 (0.81-2.20) | 0.25 | 2.00 (1.45-2.75) | <0.0001 | 1.75 (1.25-2.45) | 0.0012 |
| March | 737/4,235 (17.4) | 1.71 (1.23-2.37) | 0.0013 | 1.47 (0.87-2.46) | 0.15 | 1.71 (1.23-2.36) | 0.0014 | 1.73 (1.22-2.43) | 0.0020 |
| **Health sector** |  |  |  |  |  |  |  |  |  |
| Private | 19,763 /105,409 (18.8) | Reference |  | Reference |  | Reference |  | Reference |  |
| Public | 31,274 /113,856 (27.5) | 1.99 (1.70-2.33) | <0.0001 | 1.50 (1.28-1.75) | <0.0001 | 1.99 (1.70-2.33) | <0.0001 | 1.28 (1.12-1.48) | 0.0005 |
| **Province** |  |  |  |  |  |  |  |  |  |
| Western Cape | 9,739 /45,341 (21.5) | Reference |  | Reference |  | Reference |  | Reference |  |
| Eastern Cape | 9,334/28,671 (32.6) | 2.32 (1.79-3.01) | <0.0001 | 1.94 (1.51-2.49) | <0.0001 | 2.32 (1.79-3.01) | <0.0001 | 2.15 (1.71-2.70) | <0.0001 |
| Free State | 2,627/11,759 (22.3) | 1.14 (0.83-1.56) | 0.43 | 1.19 (0.89-1.59) | 0.24 | 1.14 (0.83-1.56) | 0.43 | 1.33 (1.01-1.74) | 0.044 |
| Gauteng | 11,938 /59,548 (20.1) | 0.76 (0.60-0.98) | 0.033 | 1.15 (0.90-1.47) | 0.28 | 0.76 (0.60-0.98) | 0.033 | 1.02 (0.82-1.26) | 0.88 |
| KwaZulu-Natal | 10,411/42,712 (24.4) | 1.45 (1.13-1.86) | 0.0040 | 1.41 (1.11-1.80) | 0.0056 | 1.45 (1.13-1.86) | 0.0040 | 1.41 (1.13-1.75) | 0.0021 |
| Limpopo | 2,431/8,110 (30.0) | 1.98 (1.44-2.72) | <0.0001 | 1.32 (0.95-1.82) | 0.095 | 1.98 (1.44-2.72) | <0.0001 | 1.72 (1.30-2.27) | 0.0001 |
| Mpumalanga | 2,209/8,287 (26.7) | 2.27 (1.62-3.20) | <0.0001 | 1.43 (0.99-2.06) | 0.060 | 2.27 (1.62-3.20) | <0.0001 | 2.01 (1.50-2.71) | <0.0001 |
| North West | 1,651/11,343 (14.6) | 0.73 (0.49-1.09) | 0.12 | 1.18 (0.79-1.76) | 0.43 | 0.73 (0.49-1.09) | 0.12 | 0.91 (0.64-1.27) | 0.57 |
| Northern Cape | 697/3,494 (20.0) | 1.32 (0.84-2.08) | 0.23 | 1.43 (0.85-2.40) | 0.18 | 1.32 (0.84-2.08) | 0.23 | 1.44 (0.97-2.14) | 0.073 |

OR = Odds Ratio, CI = 95% confidence interval
\* Chronic pulmonary disease variable excluded in the final multivariate model due to being not significant.

Sensitivity analysis of risk factors for mortality in public and private sector (Table S3, page 9 Supplementary Materials), confirm increased association of mortality for age, male sex, race and comorbidities. Divergent findings in the health sector analysis included (a) hypertension was associated with mortality in the private but not the public sector; (b) a stronger effect of age, sex, race and comorbidities in the private sector; and (c) a stronger effect of province and month of admission in the public sector, with a significant difference in mortality observed only at the peak of the second wave in the private sector.

### Association of in-hospital COVID-19 mortality with immune suppression among PLWH

When adjusting for age, sex, race, health sector, province, month of admission, NCDs, and past or active TB, compared to HIV-negative individuals, PLWH not on ART [aOR 1.45, 95% CI: 1.22-1.72] were more likely to die in-hospital compared to PLWH on ART; PLWH with history of immune suppression (CD4<200 cells/μl) [aOR 2.31, 95% CI: 1.82-2.93] were more likely to die in-hospital compared to PLWH with CD4≥200; and PLWH with history of VL≥1,000 [aOR 1.55, 95% CI: 1.20-2.01) were more likely to die in-hospital compared to PLWH with VL<1,000. (Table 3).

**Table 3:** Effect of ART, CD4 count and HIV viral load on in-hospital mortality^*^ among laboratory-confirmed SARS-CoV-2 PLWH admitted to hospital, 5 March 2020-27 March 2021, DATCOV, South Africa.

| Characteristic | Case Fatality Ratio (CFR)<br>n/N (%) unimputed | Case Fatality Ratio<br>(CFR) % (95% CI)<br>imputed | Unadjusted OR<br>(95% CI) imputed | p-value | Adjusted OR (95%<br>CI) imputed** | p-value | Adjusted OR (95%<br>CI) imputed** | p-value |
| --- | --- | --- | --- | --- | --- | --- | --- | --- |
| **ART status** |  |  |  |  |  |  |  |  |
| HIV negative | 30,697 /137,986 (22.3) | 23.1 (22.9-23.3) | Reference |  | Reference |  | 0.77 (0.72-0.82) | <0.0001 |
| HIV positive on ART | 2,046/7,484 (27.3) | 24.6 (23.8-25.4) | 0.85 (0.80-0.90) | <0.0001 | 1.30 (1.22-1.39) | <0.0001 | Reference |  |
| HIV positive not on ART | 192/594 (32.3) | 28.1 (25.2-31.1) | 0.99 (0.86-1.15) | 0.98 | 1.89 (1.60-2.23) | <0.0001 | 1.45 (1.22-1.72) | <0.0001 |
| **CD4 count** |  |  |  |  |  |  |  |  |
| HIV negative | 30,697 /137,986 (22.3) | 23.1 (22.9-23.3) | Reference |  | Reference |  | 1.06 (0.93-1.20) | 0.37 |
| HIV positive CD4≥200 | 368/1,690 (21.8) | 19.7 (17.7-21.7) | 0.64 (0.57-0.73) | <0.0001 | 0.95 (0.83-1.08) | 0.37 | Reference |  |
| HIV positive CD4<200 | 380/1,080 (35.2) | 32.2 (29.9-34.5) | 1.23 (1.09-1.38) | 0.0023 | 2.19 (1.92-2.49) | <0.0001 | 2.31 (1.82-2.93) | <0.0001 |
| **Viral load** |  |  |  |  |  |  |  |  |
| HIV negative | 30,697 /137,986 (22.3) | 23.1 (22.9-23.3) | Reference |  | Reference |  | 0.83 (0.76-0.90) | 0.0002 |
| HIV positive VL<1,000 | 316/1,273 (24.8) | 24.7 (23.0-26.4) | 0.86 (0.78-0.94) | 0.0029 | 1.21 (1.11-1.32) | 0.0002 | Reference |  |
| HIV positive VL≥1,000 | 128/443 (28.9) | 25.2 (21.0-29.4) | 0.85 (0.69-1.05) | 0.13 | 1.88 (1.53-2.31) | <0.0001 | 1.55 (1.20-2.01) | 0.0029 |

\* Model adjusted for age, sex, race, health sector, province, month of admission, non-communicable comorbidities and past or active TB
\*\*Output from the same model but with different reference categories to assess the effect of the predictors compared to HIV-uninfected individuals (model 1) or individuals not on ART, with high CD4 count, or with low viral load (model 2).

### Effect of multiple comorbidities on COVID-19 mortality

Among PLWH, the prevalence of other comorbidities was 29.2% compared to 30.8% among HIV-uninfected individuals. In a multivariable model adjusting for age, sex, race, HIV (for the non-stratified model on HIV status only), health sector, province, and month of admission, there was an increasing odds of in-hospital mortality for individuals with multiple non-HIV comorbidities, irrespective of HIV status among PLWH and HIV-uninfected individuals (Table 4). There was no statistical evidence of interaction between the presence of other multiple comorbidities and HIV infection status on in-hospital mortality (Table 4). No interaction was found for individual co-morbidities and HIV infection status on in-hospital mortality (Table S4, page 11 Supplementary Materials).

**Table 4:** Effect of single or multiple non-communicable comorbidities* on in-hospital mortality^**^ among laboratory-confirmed SARS-CoV-2 individuals admitted to hospital, 5 March 2020-27 March 2021, DATCOV, South Africa.

| Characteristic | Case fatality ratio (CFR) n/N (%) unimputed | Case fatality ratio (CFR) % (95% CI) imputed | Adjusted OR (95% CI) imputed | p-value |
| --- | --- | --- | --- | --- |
| **Any individual** |  |  |  |  |
| Comorbid condition |  |  |  |  |
| No comorbidity | 2,507/15,469 (16.2) | 15.6 (15.3-15.9) | Reference |  |
| 1 comorbid | 3,764/14,402 (26.1) | 24.1 (23.7-24.6) | 1.34 (1.28-1.39) | <0.0001 |
| 2 comorbid | 3,477/10,137 (34.3) | 31.7 (31.2-32.2) | 1.66 (1.59-1.74) | <0.0001 |
| ≥3 comorbid | 1,353/3,317 (40.8) | 41.5 (40.7-42.3) | 2.40 (2.28-2.53) | <0.0001 |
| **HIV-negative individuals** |  |  |  |  |
| Comorbid condition |  |  |  |  |
| No comorbidity | 2,074/12,993 (16.0) | 15.6 (15.3-15.9) | Reference |  |
| 1 comorbid | 3,277/12,586 (26.0) | 24.3 (23.8-24.8) | 1.34 (1.28-1.40) | <0.0001 |
| 2 comorbid | 3,082/8,962 (34.4) | 32.0 (31.4-32.6) | 1.66 (1.58-1.74) | <0.0001 |
| ≥3 comorbid | 1,159/2,858 (40.6) | 41.5 (40.4-42.5) | 2.39 (2.24-2.54) | <0.0001 |
| **HIV-positive individuals** |  |  |  |  |
| Comorbid condition |  |  |  |  |
| No comorbidity | 341/2,073 (16.5) | 15.6 (14.6-16.6) | Reference |  |
| 1 comorbid | 384/1,524 (25.2) | 23.2 (21.9-24.5) | 1.34 (1.20-1.49) | <0.0001 |
| 2 comorbid | 292/932 (31.3) | 29.4 (27.7-31.0) | 1.67 (1.48-1.90) | <0.0001 |
| ≥3 comorbid | 143/352 (40.6) | 41.9 (40.0-43.8) | 2.46 (2.22-2.73) | <0.0001 |
| **Interaction effect of multiple comorbidities with HIV infection (HIV-negative is the reference group)** |  |  |  |  |
| Comorbid condition |  |  |  |  |
| No comorbidity |  |  | Reference |  |
| 1 comorbid | N/A | N/A | 1.00 (0.89-1.12) | 0.98 |
| 2 comorbid |  |  | 1.01 (0.89-1.15) | 0.88 |
| ≥3 comorbid |  |  | 1.03 (0.90-1.17) | 0.65 |

\* Comorbid conditions included were a combination of hypertension, chronic cardiac or renal disease, diabetes, malignancy, obesity and past or active TB
\*\* Model adjusted for age, sex, race, HIV (for the non-stratified model on HIV status only), health sector, province, and month of admission

## Discussion

Among a large cohort of hospitalised individuals in a high HIV and TB prevalence setting, while age was the most important predictor of mortality, we found that HIV and TB were associated with a moderately increased risk of in-hospital COVID-19 mortality, similar to the increased risk associated with other underlying conditions such as diabetes, chronic renal disease and malignancy.

This cohort includes 13,793 PLWH and 5,282 TB patients, and in addition 2,312 patients co-infected with SARS-CoV-2, HIV and TB. Describing mortality risk amongst these groups are important as studies have demonstrated that amongst SARS-CoV-2 cases, PLWH and individuals with TB had altered T cell functions and were at risk for more severe disease. (28) Large population cohorts from the Western Cape Province of South Africa (18) and the United Kingdom (19,20) and more recent meta-analyses (21-23) found PLWH to have an increased risk of COVID-19 mortality, with increased in-hospital mortality reported in the Western Cape (18) and United Kingdom studies (19). Our study however could not conclude that HIV is a risk factor at a population level.

We describe an increased risk for in-hospital mortality for PLWH not on ART, with increasing HIV-associated immunosuppression, and with higher viral load, although missing data limited inference. Few studies have explored the association between antiretroviral treatment (ART) status, increased immunosuppression and viraemia on COVID-19 mortality among PLWH. A Western Cape public sector study found no association with the presence of high viral load or immunosuppression, using latest CD4 and viral load performed within the past 18 months through linkage with laboratory information systems. This could be explained by the fact that our study used a larger dataset over a longer time period, including data from the private sector, and from other provinces with different levels of access to care and treatment and different HIV and TB burden.

The HIV prevalence in our study was 2.2% in the private sector and 20.4% in the public sector, due to socio-economic differences in the populations served by each sector. The lower prevalence of HIV (11.7%) in individuals admitted to hospital with SARS-Cov-2 in our study, compared to the population prevalence amongst adults of 19%, is likely due to the over-representation of private sector COVID-19 admissions and possible underreporting of comorbidities in the public sector.

The prevalence of other comorbidities including TB, malignancy, chronic cardiac and renal disease, was high among PLWH included in our analysis. This high prevalence could be due to antiretroviral drugs (ARVs). For example, tenofovir disoproxil fumarate (TDF), which is part of the first line ART regimen in South Africa, has new or worsening renal failure as one of its side-effects (29) and other ARVs have side effects that include hyperlipidaemia, cardiac disease, diabetes and liver disease. (30) Also, as PLWH on ART live longer, the risk of developing NCDs increases with age. PLWH with immunosuppression are also more likely to develop TB and HIV-related malignancies. (31) Increasing numbers of comorbidities were associated with increased risk of COVID-19 mortality possibly due to poorer overall health status, more compromised immunity and presence of chronic inflammatory state, which could create a pathway for severe COVID-19 disease. (31) The effect was similar in PLWH and HIV-uninfected individuals.

Age was the strongest predictor of COVID-19 mortality as is reported in meta-analyses. (32,33) In our cohort 67% of patients who died were younger than 70 years. A review of 27 countries showed a greater proportion of deaths in LMIC occur at lower age, with people younger than 70 years constituting 63% of deaths attributed to COVID-19 in LMICs on average, versus 13% in HICs. (34) Lower recovery rates in middle-aged adults are thought to be driven by high prevalence of pre-existing conditions in younger people in LMIC, and limited access to hospitals and intensive care. (34,35) In LMIC, younger people with comorbidities should also be prioritised for vaccination along with the elderly.

While increasing age was the most important predictor, male sex, non-white race and chronic underlying illness were associated with increased mortality in our study population, as reported in meta-analyses. (2,3,5,6) For NCDs, immune function impairment, severe hypoxaemia, inflammatory activation and hypercoagulability may be contributory mechanisms for increased mortality. (36) Increased mortality associated with non-white race could be related to their burden of infection and prevalence of comorbidities. Race as a potential proxy for poverty has been shown in other studies as an additional risk factor for higher in-hospital mortality (7-9) and has been suggested to be related to structural discrimination towards marginalised populations, translating to inequities in the delivery of care and barriers to accessing care. (7)

The mortality rate of 23% observed among hospitalised patients in our analysis was at the upper bounds of the range of 15% and 24% reported in meta-analyses. (2-4) Observed variation in CFRs at different times of the epidemic and between regions and health sectors may be a result of population demographics and prevalence of comorbidities, varying population levels of COVID-19 infection, changes in admission practices, the severity of illness in admitted cases, limited access to care, higher numbers of admissions overwhelming services, improved care and treatment options as the pandemic progressed, health services’ effectiveness and completeness of death reporting. (8,37,38)

### Strengths and Limitations

The main strengths of this analysis are that the surveillance system is nationally representative across all provinces, has 100% coverage of hospitals in the public and private health sector in South Africa and contains close to 220,000 SARS-Cov-2 hospital admissions.

There were several limitations related to the study population and data completeness.

Patients with SARS-CoV-2 reported to DATCOV either had COVID-19 symptoms, were admitted for isolation, developed nosocomial COVID-19 or tested positive SARS-Cov-2 incidentally during admission for unrelated reasons. We were not able to exclude patients admitted for isolation and those with incidental findings due to incomplete data on reason for admission to DATCOV, and this may introduce bias, although likely due to high pressure for hospital beds, such individuals would have made up a small percent of included cases.

The hospitals reported a small number of 219 deaths due to cause other than COVID-19, however we have little information on the process used by the hospital for classification of cause of death other than that it is done by the attending clinician. It is possible that other deaths reported to DATCOV have not undergone similar review and may have been misclassified as COVID-19 deaths. It is also possible that some COVID-19 admissions have not been reported to the surveillance system.

Patients who were transferred to other hospitals and had no further records of admission, and those still in hospital at the time of the analysis were excluded. We do not know whether the CFR among excluded patients would differ than that of included patients possibly introducing biases. However, given the small number of the excluded patients (1.8%) the effect of biases would be limited.

There is an over-representation of private sector hospitalisations due to a lower threshold for hospitalisation in private hospitals, resulting in similar proportions of SARS-CoV-2 cases hospitalised in the public and private sector even though they serve 84% and 16% of the population respectively. This may explain why the HIV prevalence in the overall cohort (11.7%) was lower than the population prevalence of HIV in South Africa (14%). (11) However, HIV prevalence among hospitalised individuals in the public sector aged 20-59 years was similar to population HIV prevalence in this age group (Table S2, page 6 Supplementary Materials).

Data quality in a surveillance system is dependent on the information submitted by healthcare institutions. Data on comorbidities were submitted to DATCOV by the hospital based on information contained in the patient’s written or electronic hospital record, and were not independently verified as there are no systematic information systems that would verify pre-existing comorbidities. We used multiple imputation to address missing data; however, the validity of the imputed data relies on the assumption that data were missing at random. Fields with the highest proportion of incomplete data included, race (33.5%) and comorbidities (25.5% - 32.2%). The level of control of NCDs such as diabetes, using objective measures like HbA1C, was not consistently reported. Other data have shown that individuals with poorly controlled NCD were at greater risk for mortality. (39)

The most recent CD4 and VL results in the past year were submitted by the hospital based on information contained in the patient’s hospital record and not obtained from laboratory information systems. These fields had a high degree of missing data, which is an important limitation given that this analysis focuses on outcomes in PLWH. Also, the hospital did not record the date of the CD4 and VL test, so we are not able to calculate the median time between the test and COVID-19 admission, or to be certain that the status of immunosuppression or viraemia had not changed, which may also introduce measurement bias. It must be noted that DATCOV is a new surveillance system that has not yet been developed to link to other data sources and allow, for example, linkage of laboratory records to the hospitalisation record.

## Conclusions and policy implications

This study confirmed age as the most important predictor of in-hospital COVID-19 mortality in South Africa, as well as sex, race and comorbid disease. The demonstration of modest increases in COVID-19 mortality for individuals with HIV and TB (particularly PLWH who were not on ART) are important, given the high prevalence of these diseases in South Africa. PLWH, particularly those with additional comorbidities, would benefit from COVID-19 prevention programmes such as vaccine prioritisation, as well as early referral and treatment programmes that include prioritizing linkage and retention to HIV care, antiretroviral therapy adherence, virologic suppression and subsequent immune restoration. The increased CFR in the public sector, in certain provinces and during the peaks of the epidemic require further interrogation for resources and support to be directed where they are found to be required, ahead of a possible resurgence of cases.

## Data Availability

All data is available through the National Institute for Communicable Diseases by permission to the corresponding author.

## Conflicts of interest

The authors declare that there are no conflicts of interest.

## Contributorship

DS, SG, WJ contributed to literature search. WJ, MM, CC contributed to study design, data collection. WJ, PM, ST contributed to data analysis, and creation of tables and figures. WJ, ST, PM, MM verified the underlying data. WJ, ST, CC, TK, SW, LB contributed to data interpretation and writing. WJ drafted the manuscript and all other authors contributed scientific inputs equally to drafts of the manuscript.

## Acknowledgements

The authors wish to acknowledge the support and contribution from the following organisations: NICD, the National Department of Health, the nine provincial departments of health, the Hospital Association of Southern Africa (HASA), the private hospital groups and public sector hospitals who submitted data to DATCOV. Health professionals submitted data and are acknowledged and are listed as DATCOV author group at https://www.nicd.ac.za/diseases-a-z-index/covid-19/surveillance-reports/daily-hospital-surveillance-datcov-report/

The authors would also wish to acknowledge the DATCOV team, Rebone Kai, Simphiwe Dyasi, Tracy Arendse, Beverley Cowper, Kholofelo Skhosana, Felicia Malomane, Monwabisi Blom, Akhona Mzoneli, Salaminah Mhlanga, Bracha Chiger, Busisiwe Ali, Siphamandla Mzobe, Linamandla Qekeleshe, Caroline Magongwa, Caroline Mudara, Lovelyn Nnaji, Richard Welch, Noel Mfongeh, Pinky Manana, Yongama Mangwane, Mami Mokgosana, Thobile Buthelezi, Portia Makwene, Murray Dryden; the Partners in Performance team, Christo Greyling, David Spade, Winrie Kruger, Supriya Soorju and Ryan Hudson; and the ComUnity team, Gareth Jane, David Prosser, Tina Hay, Kelello Moloi, and George Ulloa.

## Disclaimer

The findings and conclusions in this report are those of the authors and do not necessarily represent the official position of the US Centers for Disease Control and Prevention, USA.

## Data sharing statement

Data used in this manuscript are available upon reasonable request. Individual participant data that underlie the results reported in this article, after de-identification (text, tables, figures, and appendices) will be shared with investigators whose proposed use of the data has been approved by an independent review committee (“learned intermediary”) identified for this purpose. Proposals should be directed to; to gain access, data requestors will need to sign a data access agreement. The request will have to be approved by the National Department of Health.

## STROBE/RECORD checklist

|  | Item | Recommendation | Reported on manuscript page |
| --- | --- | --- | --- |
| Title and abstract | 1 | (a) Indicate the study's design with a commonly used term in the title or the abstract<br>(b) Provide in the abstract an informative and balanced summary of what was done and what was found | 4 |
| Introduction |  |  |  |
| Background/rationale | 2 | Explain the scientific background and rationale for the investigation being reported | 5-6 |
| Objectives | 3 | State specific objectives, including any prespecified hypotheses | 6 |
| Methods |  |  |  |
| Study design | 4 | Present key elements of study design early in the paper | 6 |
| Setting | 5 | Describe the setting, locations, and relevant dates, including periods of recruitment, exposure, follow-up, and data collection | 6 |
| Participants | 6 | Cross-sectional study—give the eligibility criteria, and the sources and methods of selection of participants | 6 |
| Variables | 7 | Clearly define all outcomes, exposures, predictors, potential confounders, and effect modifiers. Give diagnostic criteria, if applicable | 7-8 |
| Data sources/measurement | 8* | For each variable of interest give sources of data and details of methods of assessment (measurement). Describe comparability of assessment methods if there is more than one group | 7-8 |
| Bias | 9 | Describe any efforts to address potential sources of bias | 15-17 |
| Study size | 10 | Explain how the study size was arrived at | N/A |
| Quantitative variables | 11 | Explain how quantitative variables were handled in the analyses. If applicable, describe which groupings were chosen, and why | 7-8 |
| Statistical methods | 12 | (a) Describe all statistical methods, including those used to control for confounding<br>(b) Describe any methods used to examine subgroups and interactions<br>(c) Explain how missing data were addressed<br>(d) Cross-sectional study—if applicable, describe analytical methods taking account of sampling strategy<br>(e) Describe any sensitivity analyses | 8-9<br>15-17<br>8-9<br>15-17<br>8-9<br>8-9 |
| Results |  |  |  |
| Participants | 13* | (a) Report the numbers of individuals at each stage of the study—eg, numbers potentially eligible, examined for eligibility, confirmed eligible, included in the study, completing follow-up, and analysed<br>(b) Give reasons for non-participation at each stage<br>(c) Consider use of a flow diagram | 9-10<br>Figure 1 |
| Descriptive data | 14* | (a) Give characteristics of study participants (eg, demographic, clinical, social) and information on exposures and potential confounders<br>(b) Indicate the number of participants with missing data for each variable of interest | 9-10<br>10 |
| Outcome data | 15* | Cross-sectional study—report numbers of outcome events or summary measures | 10-12 |
| Main results | 16 | (a) Give unadjusted estimates and, if applicable, confounder-adjusted estimates and their precision (eg, 95% confidence interval). Make clear which confounders were adjusted for and why they were included<br>(b) Report category boundaries when continuous variables were categorised<br>(c) If relevant, consider translating estimates of relative risk into absolute risk for a meaningful time period | 11-12 |
| Other analyses | 17 | Report other analyses done—eg, analyses of subgroups and interactions, and sensitivity analyses | 12 |
| Discussion |  |  |  |
| Key results | 18 | Summarise key results with reference to study objectives | 12-14 |
| Limitations | 19 | Discuss limitations of the study, taking into account sources of potential bias or imprecision. Discuss both direction and magnitude of any potential bias | 14-16 |
| Interpretation | 20 | Give a cautious overall interpretation of results considering objectives, limitations, multiplicity of analyses, results from similar studies, and other relevant evidence | 16 |
| Generalisability | 21 | Discuss the generalisability (external validity) of the study results | 16 |
| Other information |  |  |  |
| Funding | 22 | Give the source of funding and the role of the funders for the present study and, if applicable, for the original study on which the present article is based | 9 |

## Notes

### Competing Interest Statement

The authors have declared no competing interest.

### Funding Statement

DATCOV is funded by the National Institute for Communicable Diseases (NICD) and the South African National Government. No additional funding was obtained towards the completion of this analysis and the development of this manuscript.

### Author Declarations

The Human Research Ethics Committee (Medical), University of the Witwatersrand, approved the project protocol as part of a national surveillance program (M160667).

### Summary of Updates

Previous version used data until August 2020, this version used updated data until 27 March 2021 and utilised multiple imputation.

