## Supplementary Materials for "A national cohort study of COVID-19 in-hospital mortality in South Africa: the intersection of communicable and non-communicable chronic diseases in a high HIV prevalence setting"

Manuscript reference number: thelancethiv-D-21-00140R1

Author: Dr Waasila Jassat

Supplementary Materials

### Supplementary Methods

**DATCOV Implementation**

DATCOV was developed in March 2020 as a sentinel hospital surveillance system for COVID-19. It utilised the WHO clinical CRF to develop an online platform for data entry. The first hospitals to report on DATCOV were the large public hospitals in each province that had been designated as COVID-19 treatment facilities when the numbers of admissions were low in South Africa and treatment was limited to a few central and tertiary hospitals. The Western Cape province have an electronic information system so they began sharing all public hospital data through data export. The private hospital groups also began sharing the same. Some provinces saw the value of the surveillance system and began implementing DATCOV in more of their hospitals. By 15 July 2020, the National Department of Health (NDoH) decided to adopt DATCOV as a national hospital surveillance system for COVID-19 and provide support and resources to roll-out to all public hospitals. By October 2020, all hospitals were reporting to DATCOV. As new hospitals began to participate, they were expected to back capture all admissions since the start of the epidemic. It therefore did not matter that they had joined late as they retrospectively captured all COVID-19 admission data. The NDoH also placed data capturers at each hospital in seven provinces whose sole responsibility was to improve DATCOV data completeness and quality, and this ensured that all COVID-19 admissions were captured on DATCOV.

**Data management in DATCOV**

Data entry: DATCOV is the first electronic information system introduced to public hospitals in most provinces. There has not previously been a mandate or culture for clinicians or administrative staff to report on patient level information. When DATCOV was introduced to a few hospitals, a handful of medical doctors assumed responsibility for submitting data on the platform. During the first wave when the hospitals were busy, data capturers were required to enter data using the patient record. These data capturers may not have been able to extract all required information from the patient record, especially those related to comorbidities, complications and clinical treatment. It is also true that medical records are poorly completed in the public sector as there are no billing implications and because there are inconsistent clinical governance efforts to address this. While DATCOV was developed as an online platform for direct data entry, when it was observed that the private hospital groups and the Western Cape public sector had available data from electronic information systems, DATCOV adapted to allow for data imports. It was difficult to match all fields consistently to DATCOV so a compromise was reached and only the minimum required fields were imported (patient identifiers, age, sex, race, pregnancy, HCW status, comorbidities, and details of the admission such as date of admission, ward of care, oxygen and ventilation required and the outcome and date of outcome). The remaining variables are therefore unknown for all patients from the private sector and Western Cape public sector, which is half the patients admitted.

Data quality assurance on data entry: Most fields in DATCOV have drop-down menus or tick-boxes, and very little free text is captured. There are data validations included on DATCOV to ensure that certain data submission errors are avoided, e.g. dates not in the future, rules for minimum and maximum of weight, numerics only for certain fields, etc.

Data management: The DATCOV team do data import of private sector and Western Cape data daily, the import overwrites existing data where it has changed, e.g. outcome hand date of outcome has now been added for a patient who was in hospital. Once all imports are completed, daily linelists and reports are produced and shared with the provinces and NDoH daily by 15h00 including weekends and public holidays. While the timelines for producing reports are demanding, the DATCOV team conduct a number of routine data quality assurance steps.

Data quality assurance in data management:

1. Database validation rules during import to check for errors in incomplete data, incorrect formatting of dates, data outliers such as age older than 100.
2. Routine audit of import file and database to ensure all fields correctly imported and updated.
3. Routine audit of summary data to look for outliers, e.g. long hospital stays over 3 weeks. The hospitals are contacted and required to update if these patients have recorded outcomes.
4. Routine matching with national SARS-CoV-2 laboratory case linelist, to match against DATCOV, assign laboratory case identifier, to ensure the admission did have a recorded positive diagnosis associated with the admission. Admissions that were not found on the case linelist are investigated by the hospital, then flagged as negative and removed from reports.
5. Routine matching with other sources of data, where available, to obtain data that may be missing on DATCOV, e.g. Identity Number.

**Data collection**

DATCOV contains data on all individuals, irrespective of age, who had a positive reverse transcription polymerase chain reaction (RT-PCR) assay or antigen test for SARS-CoV-2, with a confirmed duration of stay in hospital of one full day or longer, regardless of age or reason for admission. Provinces are responsible for validating completeness of reporting on DATCOV, but this was not done systematically and it is possible that not all admissions were reported.

The surveillance system was adapted from the World Health Organization (WHO) COVID-19 case reporting tool[1], recording the following variables: demographic data, exposures such as occupation, and potential risk factors such as comorbid disease(s), and pregnancy status. Additional variables included level of treatment, complications, treatment, and outcomes of the hospital admission (discharged, transferred out to another hospital, or died).

**Time period of analysis**

[1] Global COVID-19 Clinical Data Platform for clinical characterization and management of hospitalized patients with suspected or confirmed COVID-19. Geneva: World Health Organization; 2020 Accessed 20 June 2020: https://www.who.int/teams/health-care-readiness-clinical-unit/covid-19/data-platform/.

We analysed DATCOV data from 5 March 2020 to 27 March 2021. The study period was not predefined a priori, but the endpoint was selected to provide the most updated analysis possible, and because it included all COVID-19 admissions in the first and second waves.

**Study population**

We included only the most recent admission if a patient had multiple admissions during the study period, to ensure that all admissions were independent of one another at a patient level and to avoid biasing the mortality data, as it would be impossible for a patient to die during an admission that was not their final admission in the time period. We excluded patients still in hospital and those that had been transferred out but for whom we had no follow-up record from the hospital they were transferred to. Unique patient identifiers have not been adopted in South Africa and matching re-admissions is sometimes difficult if names, surnames or identity numbers were captured differently. There are therefore a number of patients whose subsequent admissions we could not definitively match. It is highly probable that most of these transfers were of patients who were recovering from the acute COVID-19 illness and were transferred to a field hospital to continue treatment to free a bed in the acute hospital. We therefore do not believe that a large proportion of these patients would be expected to die, and we note the low CFR in field hospitals. It is however possible that amongst patients in hospital there are those with long stays because of severe disease, and amongst those transferred out there are those who are severe and transferred to higher levels of care. However, we believe that the patients excluded were probably a mix of mild and severe cases, and not dissimilar from the patients included in the analysis. They had to be excluded as they had no in-hospital outcome.

Also excluded were patients who were ascertained by the attending clinician to have died from other causes. The WHO definition of COVID-19 death excludes deaths that were clearly due to other causes.[2] We do not have detail on the method and process used by hospitals to determine the death as non-COVID. We are also unsure if all hospitals consistently utilise the same process and report non-COVID deaths. There is a possibility that other deaths have been reported that are not COVID-related and this could introduce misclassification. However we are not able to address this and have also ascertained that ISARIC and WHO analyse all deaths reported on surveillance systems, and do not make the distinction of cause of death. We do not believe this would be a very high number of deaths.

**Variables included**

Race was self-reported and included the official categories in South Africa, Black African, White, Mixed (or Coloured) and Indian.

Comorbidities included in the analysis were hypertension, diabetes, asthma, malignancy, and chronic pulmonary, cardiac and renal diseases, HIV and current and past TB. We included the comorbidities strongly suggested to be related to severe COVID-19 and did not include others such as Down Syndrome and other intellectual disabilities that have been subsequently reported to be associated with higher mortality. We included past TB within the previous 10 years, because it has been suggested that lung damage from previous TB could result in more severe pulmonary disease with COVID-19 (https://theunion.org/our-work/covid-19/covid-19-and-tb-frequently-asked-questions).

[2] International guidelines for certification and classification (coding) of COVID-19 as cause of death. Based on ICD International Statistical Classification of Diseases (16 April 2020) https://www.who.int/classifications/icd/Guidelines_Cause_of_Death_COVID-19.pdf

Obesity was not consistently recorded, and was either based on the calculation of body mass index or on the subjective opinion of the attending healthcare worker, which could result in misclassification. There were also 75% of records with missing Obesity data. We therefore excluded Obesity from the analysis.

Data on comorbidities including HIV, ART status, CD4 and VL were entered on DATCOV by the hospital, based on information contained in the patient's written or electronic hospital record, and were not independently verified by the DATCOV team. The laboratory results were also not obtained from the laboratory information systems to the hospital record, due in part to lack of access to private laboratory data and to difficulties linking records as South Africa has not adopted a unique patient identifier. Also, the hospital did not record the date of the CD4 and VL test, so we are not able to calculate the median time between the test and COVID-19 admission, or to be certain that the status of immunosuppression or viraemia had not changed, which may also introduce measurement bias. It must be noted that DATCOV is a new surveillance system that has not yet been developed to link to other data sources and allow, for example, linkage of laboratory records to the hospitalisation record.

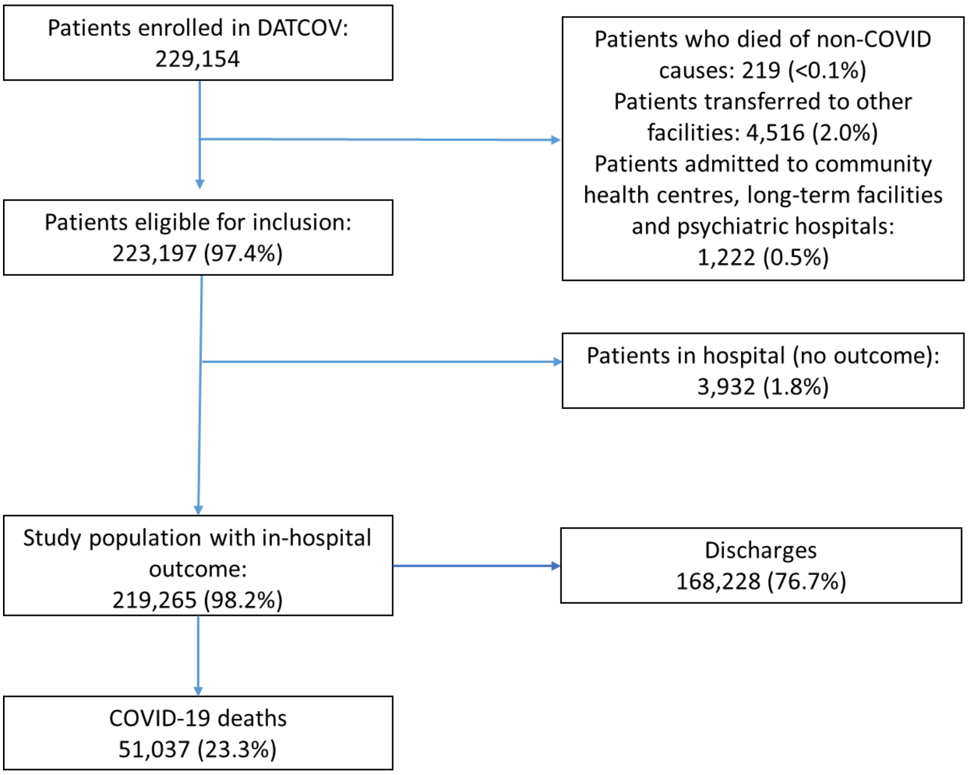


**Figure S1**: Flow diagram of cohort with numbers excluded at different stages and identification of cases for the main endpoints, 5 March 2020-27 March 2021, DATCOV, South Africa.

**Table S1:** Missing data and characteristics of COVID-19 hospitalised patients reported to DATCOV, 5 March 2020-27 March 2021, DATCOV, South Africa.

| Characteristic | Missing data (N=219,265) n (%) | Unimputed n (%) | Imputed % (95% CI) |
|---|---|---|---|
| **Sex** | **438 (0.2)** | **N=218,827** | **N=219,265** |
| Female | | 121,937 (55.7) | 55.7 (55.5-55.9) |
| Male | | 96,890 (44.3) | 45.6 (45.2-46.1) |
| **Age (in years)** | **Complete variable** | **N=219,265** | **N=219,265** |
| <20 | | 8,605 (3.9) | 3.9 (3.8-4.0) |
| 20-30 | | 44,605 (20.3) | 20.3 (20.2-20.5) |
| 40-59 | | 86,095 (39.3) | 39.3 (39.1-39.5) |
| 60-69 | | 41,077 (18.7) | 18.7 (18.6-18.9) |
| 70-79 | | 24,943 (11.4) | 11.4 (11.2-11.5) |
| ≥80 | | 13,940 (6.4) | 6.4 (6.3-6.5) |
| **Race** | **73,389 (33.5)** | **N=145,876** | **N=219,265** |
| White | | 12,661 (8.7) | 10.9 (10.7-11.1) |
| Mixed | | 10,053 (6.9) | 11.2 (11.0-11.4) |
| Black | | 114,571 (78.5) | 72.2 (71.9-72.5) |
| Indian | | 8,591 (5.9) | 5.7 (5.6-5.8) |
| **Hypertension** | **55,915 (25.5)** | **N=163,350** | **N=219,265** |
| No | | 102,252 (62.6) | 61.4 (61.1-61.6) |
| Yes | | 61,098 (37.4) | 38.6 (38.4-38.9) |
| **Chronic cardiac disease** | **69,233 (31.6)** | **N=150,032** | **N=219,265** |
| No | | 145,854 (97.2) | 95.1 (94.9-95.2) |
| Yes | | 4,178 (2.8) | 4.9 (4.8-5.1) |
| **Chronic lung disease/Asthma** | **67,039 (30.6)** | **N=152,226** | **N=219,265** |
| No | | 141,726 (93.1) | 92.2 (92.0-92.3) |
| Yes | | 10,500 (6.9) | 7.8 (7.7-8.0) |
| **Chronic kidney disease** | **70,049 (31.9)** | **N=149,216** | **N=219,265** |
| No | | 145,095 (97.2) | 95.9 (95.7-96.1) |
| Yes | | 4,121 (2.8) | 4.1 (3.9-4.3) |
| **Diabetes** | **59,333 (27.1)** | **N=159,932** | **N=219,265** |
| No | | 116,047 (72.6) | 71.0 (70.8-71.2) |
| Yes | | 43,885 (27.4) | 29.0 (28.8-29.2) |
| **Malignancy** | **70,463 (32.1)** | **N=148,802** | **N=219,265** |
| No | | 147,659 (99.2) | 98.9 (98.8-99.0) |
| Yes | | 1,143 (0.8) | 1.1 (1.0-1.2) |
| **Obesity** | **163,570 (74.6)** | **N=55,695** | **N=219,265** |
| No | | 49,604 (89.1) | 88.3 (87.9-88.7) |
| Yes | | 6,091 (10.9) | 11.7 (11.3-12.1) |
| **Tuberculosis** | **72,884 (33.2)** | **N=146,381** | **N=219,265** |
| Never | | 141,099 (96.4) | 94.9 (94.8-95.1) |
| Past | | 3,001 (2.1) | 2.5 (2.4-2.6) |
| Active | | 993 (0.7) | 1.0 (0.9-1.1) |
| Past and active | | 1,288 (0.9) | 1.6 (1.5-1.6) |
| **HIV** | **67,486 (30.8)** | **N=151,779** | **N=219,265** |
| No | | 137,986 (90.9) | 88.3 (88.0-88.5) |
| Yes | | 13,793 (9.1) | 11.7 (11.5-12.0) |
| **ART** | **5,715 (41.4)*** | **N=8,078** | **N=25,578** |
| On ART | | 7,484 (92.7) | 92.8 (92.5-93.1) |

| | | | |
|---|---|---|---|
| Not on ART | | 594 (7.3) | 7.2 (6.9-7.5) |
| **Viral load (RNA copies/ml)** | **12,077 (87.6)*** | **N=1,716** | **N=25,578** |
| <1,000 | | 1,273 (74.2) | 72.8 (72.3-73.4) |
| ≥1,000 | | 443 (25.8) | 27.2 (26.6-27.7) |
| **CD4 count (cells/µL)** | **11,023 (79.9)*** | **N=2,770** | **N=25,578** |
| ≥200 (immune reconstituted) | | 1,690 (61.0) | 57.9 (57.3-58.5) |
| <200 (immunosuppressed) | | 1,080 (39.0) | 42.1 (41.5-42.7) |
| **Health sector** | **Complete variable** | **N=219,265** | **N=219,265** |
| Private | | 105,409 (48.1) | 48.1 (47.9-48.3) |
| Public | | 113,856 (51.9) | 51.9 (51.7-52.1) |
| **Province** | **Complete variable** | **N=219,265** | **N=219,265** |
| Eastern Cape | | 28,671 (13.1) | 13.1 (12.9-13.2) |
| Free State | | 11,759 (5.4) | 5.4 (5.3-5.5) |
| Gauteng | | 59,548 (27.2) | 27.2 (27.0-27.3) |
| KwaZulu-Natal | | 42,712 (19.5) | 19.5 (19.3-19.6) |
| Limpopo | | 8,110 (3.7) | 3.7 (3.6-3.8) |
| Mpumalanga | | 8,287 (3.8) | 3.8 (3.7-3.9) |
| North West | | 11,343 (5.2) | 5.2 (5.1-5.3) |
| Northern Cape | | 3,494 (1.6) | 1.6 (1.5-1.6) |
| Western Cape | | 45,341 (20.7) | 20.7 (20.5-20.8) |
| **Month of admission** | **51 (<0.1)** | **N=219,214** | **N=219,265** |
| March | | 400 (0.2) | 0.2 (0.2-1.2) |
| April | | 1,449 (0.7) | 0.7 (0.6-0.7) |
| May | | 5,787 (2.6) | 2.6 (2.6-2.7) |
| June | | 18,209 (8.3) | 8.3 (8.2-8.4) |
| July | | 38,226 (17.4) | 17.4 (17.3-17.6) |
| August | | 19,671 (9.0) | 9.0 (8.9-9.1) |
| September | | 8,851 (4.0) | 4.0 (4.0-4.1) |
| October | | 7,735 (3.5) | 3.5 (3.5-3.6) |
| November | | 11,110 (5.1) | 5.1 (5.0-5.2) |
| December | | 39,582 (18.1) | 18.1 (17.9-18.2) |
| January | | 52,019 (23.7) | 23.7 (23.6-23.9) |
| February | | 11,940 (5.5) | 5.4 (5.4-5.5) |
| March | | 4.235 (1.9) | 1.9 (1.9-2.0) |
| **In-hospital outcome** | **Complete variable** | **N=219,265** | **N=219,265** |
| Discharged alive | | 168,228 (76.7) | 76.7 (76.5-76.9) |
| Died in hospital | | 51,037 (23.3) | 23.3 (23.1-23.5) |

* Number and percentage of missing data calculated among g13,793 known HIV-positive individuals.

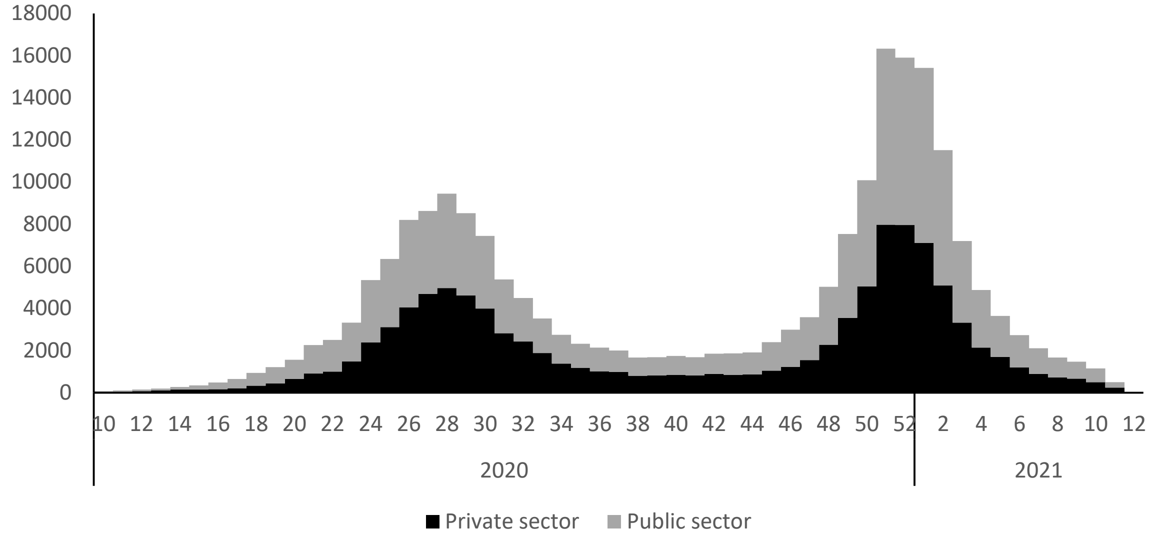


**Figure S2**: Number of reported COVID-19 admissions by health sector and epidemiological week of diagnosis among 380 hospitals, 5 March 2020-27 March 2021, DATCOV, South Africa (n=219,265)

Note: right weeks censored as only individuals with in-hospital outcome were included

**Table S2**: HIV prevalence amongst COVID-19 hospitalized patients of different age groups reported to DATCOV, in the public and private health sectors, 5 March 2020-27 March 2021, South Africa.

| Characteristic | Private sector (94,167) n/N (%) | Public sector (57,612) n/N (%) |
|---|---|---|
| **Age** | | |
| <20 years | 16/2,806 (0.6) | 211/2,261 (9.3) |
| 20-39 years | 474/16,501 (2.9) | 4,142/12,332 (33.6) |
| 40-59 years | 1,335/41,973 (3.2) | 5,516/20,556 (26.8) |
| 60-69 years | 198/16,944 (1.2) | 1,376/12,013 (11.5) |
| 70-79 years | 32/10,150 (0.3) | 403/7,337 (5.5) |
| ≥80 years | 2/5,793 (0.03) | 88/3,113 (2.8) |
| Total | 2,057/94,167 (2.2) | 11,736/57,612 (20.4) |

**Table S3**: Factors associated with in-hospital mortality among individuals with laboratory-confirmed SARS-CoV-2 admitted to hospital, by health sector, 5 March 2020-27 March 2021, DATCOV, South Africa. (N=219,265)

| Characteristic | Public sector Adjusted OR (95% CI) imputed | p-value | Private sector Adjusted OR (95% CI) imputed | p-value |
|---|---|---|---|---|
| **Sex** | | | | |
| Female | Reference | | Reference | |
| Male | 1.26 (1.22-1.30) | <0.001 | 1.33 (1.28-1.38) | <0.001 |
| **Age** | | | | |
| <20 years | Reference | | Reference | |
| 20-39 years | 1.83 (1.60-2.10) | <0.001 | 4.80 (3.40-6.78) | <0.001 |
| 40-59 years | 4.30 (3.76-4.91) | <0.001 | 13.80 (9.82-19.39) | <0.001 |
| 60-69 years | 9.00 (7.88-10.29) | <0.001 | 29.77 (21.16-41.88) | <0.001 |
| 70-79 years | 12.16 (10.62-13.92) | <0.001 | 44.93 (31.89-63.31) | <0.001 |
| ≥80 years | 13.80 (11.99-15.87) | <0.001 | 68.27 (48.33-96.44) | <0.001 |
| **Race** | | | | |
| White | Reference | | Reference | |
| Black | 1.18 (1.03-1.35) | 0.015 | 1.36 (1.22-1.50) | <0.001 |
| Mixed | 1.18 (1.06-1.32) | 0.003 | 1.34 (1.23-1.46) | <0.001 |
| Indian | 1.41 (1.19-1.67) | <0.001 | 1.39 (1.27-1.51) | <0.001 |
| **Hypertension** | | | | |
| No | | | Reference | |
| Yes | | | 1.15 (1.10-1.20) | <0.001 |
| **Diabetes** | | | | |
| No | Reference | | Reference | |
| Yes | 1.38 (1.33-1.43) | <0.001 | 1.44 (1.38-1.51) | <0.001 |
| **Chronic cardiac** | | | | |
| No | Reference | | Reference | |
| Yes | 2.45 (2.27-2.64) | <0.001 | 1.66 (1.48-1.87) | <0.001 |
| **Chronic renal disease** | | | | |
| No | Reference | | Reference | |
| Yes | 1.43 (1.30-1.58) | <0.001 | 2.06 (1.75-2.43) | <0.001 |
| **Malignancy** | | | | |
| No | Reference | | Reference | |
| Yes | 1.30 (1.11-1.53) | 0.002 | 1.98 (1.63-2.40) | <0.001 |
| **Tuberculosis** | | | | |
| None | Reference | | Reference | |
| Past | 1.22 (1.11-1.33) | <0.001 | 4.18 (2.20-7.97) | <0.001 |
| Current | 1.37 (1.14-1.64) | 0.001 | 1.52 (1.20-1.92) | <0.001 |
| Current and past | 1.44 (1.28-1.61) | <0.001 | 2.00 (0.68-5.83) | 0.206 |
| **HIV** | | | | |
| No | Reference | | Reference | |
| Yes | 1.27 (1.19-1.36) | <0.001 | 1.51 (1.34-1.71) | <0.001 |
| **Month of admission** | | | | |
| March | Reference | | Reference | |
| April | 1.75 (1.00-3.06) | 0.050 | 0.83 (0.49-1.40) | 0.489 |
| May | 2.84 (1.69-4.77) | <0.001 | 1.01 (0.63-1.62) | 0.981 |
| June | 3.38 (2.02-5.66) | <0.001 | 1.06 (0.67-1.68) | 0.811 |
| July | 3.64 (2.18-6.08) | <0.001 | 1.08 (0.69-1.71) | 0.729 |
| August | 2.75 (1.64-4.61) | <0.001 | 0.88 (0.55-1.39) | 0.577 |

| | | | | |
|---|---|---|---|---|
| September | 2.34 (1.39-3.93) | 0.001 | 0.66 (0.42-1.06) | 0.085 |
| October | 2.29 (1.36-3.85) | 0.002 | 0.69 (0.43-1.10) | 0.115 |
| November | 3.51 (2.09-5.87) | <0.001 | 1.05 (0.66-1.68) | 0.823 |
| December | 4.36 (2.61-7.29) | <0.001 | 1.66 (1.05-2.63) | 0.029 |
| January | 4.20 (2.52-7.02) | <0.001 | 1.80 (1.14-2.85) | 0.012 |
| February | 2.96 (1.77-4.96) | <0.001 | 1.05 (0.66-1.67) | 0.839 |
| March | 3.46 (2.05-5.84) | <0.001 | 0.76 (0.47-1.23) | 0.268 |
| **Province** | | | | |
| Western Cape | Reference | | Reference | |
| Eastern Cape | 2.28 (1.64-3.17) | <0.001 | 1.79 (1.41-2.28) | <0.001 |
| Free State | 1.32 (0.87-2.01) | 0.194 | 1.43 (1.12-1.82) | 0.004 |
| Gauteng | 0.86 (0.57-1.28) | 0.450 | 1.14 (0.97-1.33) | 0.113 |
| KwaZulu-Natal | 1.52 (1.08-2.15) | 0.017 | 1.28 (1.07-1.52) | 0.007 |
| Limpopo | 1.85 (1.25-2.75) | 0.002 | 1.55 (1.13-2.13) | 0.006 |
| Mpumalanga | 2.53 (1.64-3.90) | <0.001 | 1.23 (0.91-1.65) | 0.178 |
| North West | 0.87 (0.50-1.52) | 0.636 | 0.97 (0.74-1.26) | 0.796 |
| Northern Cape | 1.64 (0.93-2.91) | 0.088 | 1.29 (0.87-1.89) | 0.202 |

OR = Odds Ratio, CI = 95% confidence interval

**Table S4**: Effect of individual non-communicable comorbidities on in-hospital mortality** among PLWH with laboratory-confirmed SARS-CoV-2 admitted to hospital, 5 March 2020-27 March 2021, DATCOV, South Africa.

| Characteristic | Adjusted OR (95% CI) imputed | p-value |
|---|---|---|
| **HIV-negative individuals** | | |
| Hypertension | 1.06 (1.02-1.09) | 0.002 |
| Diabetes | 1.38 (1.34-1.43) | <0.001 |
| Chronic cardiac disease | 2.11 (2.00-2.23) | <0.001 |
| Chronic renal disease | 1.42 (1.31-1.54) | <0.001 |
| Malignancy | 1.54 (1.37-1.73) | <0.001 |
| Past TB | 1.33 (1.20-1.48) | <0.001 |
| Active TB | 1.28 (1.08-1.51) | 0.005 |
| Past and active TB | 1.24 (1.00-1.55) | 0.049 |
| **HIV-positive individuals** | | |
| Hypertension | 0.93 (0.84-1.03) | 0.146 |
| Diabetes | 1.29 (1.18-1.41) | <0.001 |
| Chronic cardiac disease | 2.38 (1.95-2.89) | <0.001 |
| Chronic renal disease | 1.41 (1.19-1.67) | <0.001 |
| Malignancy | 1.30 (0.96-1.75) | 0.085 |
| Past TB | 1.18 (1.03-1.35) | 0.017 |
| Active TB | 1.65 (1.25-2.18) | 0.001 |
| Past and active TB | 1.59 (1.37-1.84) | <0.001 |
| **Interaction effect of individual comorbidities with HIV infection (HIV-negative is the reference group)** | | |
| Hypertension | 0.88 (0.80-0.97) | 0.008 |
| Diabetes | 0.93 (0.85-1.03) | 0.151 |
| Chronic cardiac disease | 1.13 (0.92-1.37) | 0.226 |
| Chronic renal disease | 0.99 (0.85-1.16) | 0.901 |
| Malignancy | 0.84 (0.62-1.14) | 0.257 |
| Past TB | 0.89 (0.76-1.03) | 0.122 |
| Active TB | 1.29 (0.92-1.82) | 0.139 |
| Past and active TB | 1.28 (0.98-1.67) | 0.071 |

* Model adjusted for age, sex, race, HIV (for the non-stratified model on HIV status only), health sector, province, and month of admission
